# Cardiac Cross-Reactivity of NaV Autoantibodies in Metastatic Breast Cancer: A Possible Trigger for Sudden Cardiac Death

**DOI:** 10.1101/2024.09.06.24313111

**Authors:** Carlo Pappone, Adriana Tarantino, Dario Melgari, Marco Piccoli, Giuseppe Ciconte, Anthony Frosio, Emanuele Micaglio, Serena Calamaio, Chiara Vantellino, Federica Cirillo, Pasquale Creo, Valerio Castoldi, Rachele Prevostini, Alessandro De Toma, Antonio Boccellino, Gabriele Negro, Luigi Giannelli, Žarko Ćalović, Letizia Leocani, Vincenzo Santinelli, Domenico De Toma, Ilaria Rivolta, Luigi Anastasia

## Abstract

**Background and Aims:** Patients with metastatic breast cancer have an increased risk of sudden cardiac death (SCD) that cannot be fully explained by cardiotoxic treatments. Recent evidence shows that autoantibodies targeting the cardiac NaV1.5 sodium channel in Brugada syndrome (BrS) can trigger arrhythmias and elevate SCD risk. Similarly, autoantibodies against the neonatal NaV1.5 isoform have been found in metastatic breast cancer patients. Given the high homology between these NaV1.5 isoforms, we investigated whether these autoantibodies cross-react with the cardiac isoform, potentially contributing to SCD in this population.

**Methods:** Plasma from twenty metastatic breast cancer patients was analyzed for anti-NaV1.5 autoantibodies using HEK293A cells expressing the NaV1.5 protein, followed by Western blotting. The effects of these autoantibodies on sodium current density were assessed in cellular models and wild-type mice, with electrocardiographic monitoring after plasma infusion.

**Results:** Fifteen plasma samples from metastatic breast cancer patients tested positive for anti-NaV1.5 autoantibodies, significantly reducing sodium current density in vitro. Mice injected with these plasma samples developed severe arrhythmias and a Brugada syndrome-like ECG pattern. In contrast, plasma samples either without the autoantibodies or with IgG depletion showed no such effects, underscoring the role of IgG in sodium current reduction and confirming the pathogenicity of the autoantibodies.

**Conclusions:** This study demonstrates that anti-NaV1.5 autoantibodies in metastatic breast cancer patients can cross-react with the cardiac NaV1.5 isoform, potentially leading to fatal arrhythmias. These findings highlight a novel mechanism for the high SCD rate in this population and suggest that therapies involving sodium blockers should be used with caution to avoid exacerbating this risk. Reliable diagnostic tests and targeted therapies are urgently needed to mitigate SCD risk in affected patients.

## Introduction

Sudden cardiac death (SCD) is surprisingly common in cancer patients, often occurring unexpectedly and remaining largely unexplained^1–4^. In advanced cancer patients, the SCD incidence ranges from 6% to 20% within the first six months of treatment^2,5^. This high frequency cannot be solely attributed to the cardiotoxic effects of treatments like chemotherapy or metastases^2,5^, highlighting the need to identify new biomarkers to predict and prevent these fatal events.

Recent evidence suggests that cancer and cardiovascular disease share common molecular mechanisms, including the dysregulation of voltage-gated sodium channels (VGSCs)^6,7^. Among these channels, the cardiac NaV1.5 isoform plays a critical role in maintaining electrical stability^8,9^. Mutations in NaV1.5 are linked to a range of cardiac arrhythmias, with loss-of-function mutations being particularly associated with Brugada Syndrome (BrS). This condition is characterized by reduced sodium current and a heightened risk of ventricular fibrillation^8–11^. At the same time, NaV1.5 has also been shown to be overexpressed in several metastatic cancers, including colorectal, prostate, ovarian and breast cancers, where mainly its ‘*neonatal*’ splice variant (nNaV1.5) promotes cell motility and invasiveness through mechanisms such as increased sodium influx and subsequent degradation of the extracellular matrix^12–15^. Therapies targeting the neonatal isoform are currently being developed to suppress cancer progression without affecting other NaV isoforms, particularly the ‘*adult*’ isoform (aNaV1.5) in the heart, so that cardiac function is not compromised^14,16–21^. In this context, it has recently been reported that patients with metastatic breast cancer produce autoantibodies against nNaV1.5 as an immune response to the cancer^22^. However, the specificity of these autoantibodies and the potential for off-target effects due to molecular mimicry have not been fully investigated^9,15,22^. Additionally, it was recently discovered that patients with BrS produce autoantibodies against the cardiac NaV1.5 isoform, which alone can induce typical and lethal arrhythmias when injected into wild-type mice, indicating a possible pathogenic role^23^. These autoantibodies have also been shown to cross-react with other NaV isoforms, such as NaV1.4 in skeletal muscle^23^.

On this basis, in this study we investigated whether the circulating autoantibodies against NaV1.5 present in metastatic breast cancer patients could cross-react with the adult cardiac isoform NaV1.5 and possibly contribute to an increased risk of SCD.

## Methods

### Human samples

Twenty plasma samples from patients with metastatic breast cancer (MBC) (age 54.10±13.32) were all referred to the Arrhythmology Department of the IRCCS Policlinico San Donato and included in the BrS registry (NCT 02641431; NCT03106701)^24^. Six plasma samples from healthy subjects, who tested negative for BrS according to the current guidelines^25^, were also obtained from IRCCS Policlinico San Donato Milan, and were used as negative controls for the immunoprecipitation and FACS analyses. Inclusion criteria for breast cancer patients comprised: i) early-invasive or advanced stage breast cancer, ii) having received treatments, iii) no chronic diseases, iv) a normal ECG with no signs or history of BrS. Healthy participants had: i) no history of breast cancer, and ii) no chronic diseases, iii) a negative BrS ajmaline provocation test^25^. These samples were anonymized before analysis, and all participants provided written informed consent in accordance with the Declaration of Helsinki. The protocol of this study was reviewed and approved by the Ospedale San Raffaele Ethics Committee. The authors had full access to all data and take full responsibility for its integrity and analysis.

### Plasma Collection and IgG depletion

Blood samples (10 ml) were centrifuged at 1000 x g for 15 minutes to separate plasma. A second centrifugation at 2000 x g for 15 minutes further clarified the plasma, which was then aliquoted and stored at −20°C. For IgG depletion, 200 μl of plasma from MBC patients were incubated with Protein G magnetic beads (Thermo Fisher Scientific) for 1.5 hours at room temperature in a rotator. The procedure was repeated three times with fresh beads.

### Generation of Stable Cell Line and Transient Transfections

HEK293A cells were cultured in Dulbecco’s Modified Eagle Medium (DMEM, Life Technologies) supplemented with 10% fetal bovine serum (FBS, Sigma), 2 mM glutamine (Merck), and 1X penicillin/streptomycin (Euroclone), at 37°C in a humidified atmosphere of 5% CO_2_ and 95% air. The human NaV1.5 cDNA adult isoform was cloned into the pcDNA 3.1(+) vector. A stable cell line was obtained using jetPRIME (Euroclone), following the manufacturer’s protocol, and pharmacological selection using G418 for Western blot. For transient transfection, JetPRIME reagent was used to transfect HEK293A cells with 0.5 µg of either the adult or neonatal isoform of SCN5A cDNA (this last was a kind gift of Prof Mirko Baruscotti, University of Milano) and 0.2 µg of green fluorescent protein (eGFP) cDNA, for patch clamp experiments.48 hours after the transfection cells were detached and re-plated at single cell confluence to perform electrophysiological recordings. NaV1.5-GFP cDNA was transiently transfected in HEK293A cells using JetPRIME reagent, and after 24h were used for flow cytometry validation.

### NaV1.5 Immunoblotting

HEK293A cells overexpressing NaV1.5 were lysed using RIPA buffer containing a cocktail of protease and phosphatase inhibitors, with subsequent centrifugation at 15,000 rpm for 10 minutes at 4°C to collect the supernatant. Protein concentrations were determined by the BCA assay (Pierce). Proteins were denatured and reduced with Laemmli buffer containing β-mercaptoethanol (Bio-Rad) and loaded onto a 10% SDS-PAGE gel (Protean Tgx Stain-Free, Bio-Rad) for electrophoresis, followed by transfer to nitrocellulose membranes. The membranes were blocked and probed overnight at 4°C with the primary anti-NaV1.5 antibody (1:2000 dilution, Cell Signaling, clone D9J7S), followed by washing and incubation with a secondary anti-rabbit IRDye680 antibody (1:2000 dilution, LI-COR Biosciences). Membranes were then incubated with MBC patient plasma, followed by staining with a secondary anti-human IgG-HRP antibody (1:800 dilution, Bio-Rad). Bands were visualized using the ECL Advance kit (GE Healthcare) and imaged with the ChemiDoc MP system (Bio-Rad).

### NaV1.5 Immunoprecipitation

HEK293A cells expressing NaV1.5 were lysed as described in the Methods. MBC patient IgGs, extracted from plasma using Dynabeads™ Protein G (Thermo Fisher Scientific) according to the datasheet, were incubated with 500 μg of protein lysate for 1 hour at room temperature with orbital shaking. The beads were washed three times with TBS, and Laemmli-β-mercaptoethanol buffer was added for a 5-minute incubation at 100°C. The eluted fraction was subsequently subjected to Western blot for NaV1.5 (diluted 1:2000) overnight at 4°C, and the secondary antibody anti-rabbit IRDye 680 (diluted 1:2000) for 1 hour at room temperature.

### Flow Cytometry Assays to Detect NaV1.5-IgG

2 × 10^5^ NaV1.5-GFP transfected cells were harvested, washed twice with cold PBS pH 7.4, and incubated for 1h at 4°C with four patient plasma (three with NaV IgGs and one without) and three control plasma without NaV IgGs (diluted 1:30). Thereafter, the cells were washed, resuspended in PBS, and subjected to flow cytometry analysis using a Cytoflex S (Beckman Coulter). The optimal data acquisition gate was set for analysis, and binding was measured as the mean fluorescence intensity (MFI). NaV1.5-IgG titers were determined by the difference in MFI (ΔMFI) between NaV1.5-GFP-expressing cells and untransfected cells, as previously validated^26^.

### MTT assay cell viability

A colorimetric assay based on tetrazolium salt MTT (3-(4,5-dimethylthiazol-2-yl)-2,5-diphenyl tetrazolium bromide), was performed to evaluate the viability of cells upon the plasma treatment. Briefly, HEK293A cells expressing NaV1.5 channel, MCF-7, and MDA-MB-231 cells were cultured and exposed to 1, 5 and 10% of pre-heated plasma in transparent/clear flat bottom 96-wells plates for 1 hour followed by incubation with 0.5 mg/ml MTT for 1 hour at 37°C and 5% (v/v) CO_2_. Then the medium was removed and formed formazan crystals were dissolved in DMSO and ethanol (1:1 diluted) on a microplate shaker for 20 minutes at room temperature. Absorption was measured at 570 nm and subtracted from background at 670 nm.

### hiPSC culture and cardiac differentiation

The TMOi001-A hiPSC line from a healthy female donor was obtained from Thermo Fisher Scientific and maintained in TeSR-E8 TM medium (Thermo Fisher Scientific) on human Biolaminin 521 LN-coated dishes^27^. Cardiac differentiation was induced with the PSC Cardiomyocytes Differentiation Kit (Thermo Fisher Scientific) on monolay culture on Matrigel® hESC-qualified Matrix (Corning). After 21 days of differentiation, the PSC-Derived Cardiomyocytes Isolation Kit (Milteny Biotech) was used to enrich hiPS-CMs cultures, that were then cultured in Cardiomyocytes Maintenance Medium (CMM) with B27 supplement (Thermo Fisher Scientific) until day 28. For electrophysiological experiments, at least 48 hours before the experimental day, hiPS-CMs were detached, and single cells were plated on Matrigel-coated 35 mm dishes (VWR).

### Tissue Preparation and Immunostaining of Mouse Tissue

Tissue sections of the left ventricle from adult C57BL/6n mice were collected in our laboratory immediately after the ECG records and stored at −20 to −80°C. Then, they were cut with a cryostat (Leica CM1900), mounted on gelatin-coated histologic slides, and stored at −20 to −80°C until use. Slides were thawed at room temperature for 10-20 minutes, rehydrated in wash buffer for 10 minutes, and incubated in an antigen unmasking solution (0.01 M citrate buffer, pH 6) for 15 minutes at 98°C. To block non-specific binding, sections were incubated for 1 hour at room temperature with 5% normal donkey serum (NDS) and 5% bovine serum albumin (BSA) in PBS with 0.1% Tween 20. After washing, sections were incubated for 1 hour at room temperature with anti-human IgG-FITC antibody, diluted 1:200 in PBS containing 2% NDS and 2% BSA. After several washing steps, mounted with Vectashield mounting medium containing DAPI. Images were captured using the Leica Thunder (×40 objective).

### Sodium current recording

Sodium current (I_Na_) was recorded on hiPSCs-CM and on transfected HEK293A cells incubated with 5% plasma from MBC patients, complete or depleted of IgG, for 1 hour at 37°C and 5% CO_2_. Patch-clamp experiments were conducted in whole cell on a manual set-up mounting a 700B operational amplifier (Molecular Devices). Borosilicate glass patch pipettes were pulled with a P1000 horizontal puller (Sutter) to a resistance of 3-5 MΩ. In both the cell models, I_Na_ was elicited by a 10 mV-step protocol ranging from −80 to 60 mV. For hiPS-CMs, voltage step duration was 150 ms from a holding potential of −80 mV, while for HEK293A cells voltage step duration was 50 ms from a holding potential of −120 mV and P/4 leak subtraction was applied. All signals were sampled at 50 kHz and low pass filtered at 10 kHz. Membrane capacitance and 60-80% series resistance were compensated. All recording were performed at room temperature. The extracellular recording solution for hiPS-CMs contained (in mM): NaCl 80, N-methyl-D-glucosamine (NMDG) 60, KCl 4, CaCl_2_ 2, MgCl_2_ 1, HEPES 10, Glucose 5 (pH 7.4 HCl), 10 µM nifedipine was added to remove the L-type calcium current, and 30 µM tetrodotoxin (TTX) pharmacologically isolate the I_Na_. The intracellular pipette solution contained (in mM): CsCl 135, NaCl 10, EGTA 5, CaCl_2_ 2, TEA-Cl 2, HEPES 10, MgATP 2 (pH 7.2 CsOH). In HEK293A cells recordings, the bath solution contained (in mM): NaCl 60, NMDG 80, KCl 4, CaCl_2_ 2, MgCl_2_ 1, HEPES 10, Glucose 5 (pH 7.4 HCl). The intracellular pipette solution contained (in mM): CsF 110, CsCl 10, EGTA 10, HEPES 10 (pH 7.2 CsOH). Raw recordings were analyzed with Clampfit 10.7 (Molecular Devices), Origin Pro (OriginLab) and GraphPad Prism (GraphPad Software). For each cell, current density was calculated by dividing the current amplitude by the measured cell capacitance. The current-voltage (I-V) relationship was built by plotting the peak current density against the respective test voltage level. Steady-state activation curves were constructed from the I-V relationship and fitted with a Boltzmann sigmoidal function y = 1/(1+exp[(Vm-V1/2)/k]), where y is the relative current, Vm is the membrane potential, V1/2 is the half-maximal activation voltage, and k is the slope factor of the curve.

### Animals and Electrocardiography

The procedure involving mice was performed according to the animal protocol guidelines described by the Institutional Animal Care and Use Committee (IACUC) authorization no. 425/2022/PR at San Raffaele Scientific Institute (Milan, Italy). Mice C57BL/6n at 50 weeks were maintained *ad libitum* access to water and standard chow food at room temperature with a 12-hours light/dark schedule. They were anesthetized by intraperitoneal injection of medetomidine, 0.5 mg/kg (Orion Pharma S.r.l.) and ketamine, 100 mg/kg (Merial), both diluted in saline solution. Body weight was determined prior to each investigation, the body temperature was constantly monitored and kept at 37 ± 0.5°C by a homeothermic blanket system with a rectal thermometer probe (Harvard Apparatus, Holliston, Massachusetts, USA). 150 μl of plasma from breast cancer patients (n=4) were pre-heated at 56°C for 20 minutes before the intravenous injection in anesthetized mice. Briefly, ECG was performed continuously from 5 minutes after induction of anesthesia and 20 minutes after plasma injection using four subcutaneous needle electrodes (stainless steel, 27-gauge, 12 mm length; SEI EMG S.r.l., Cittadella, Italy): two needles were inserted in the forelimbs and one in the left hindlimb, and another needle electrode was placed in the right hindlimb as a ground. A consistent lead configuration without changing the polarity or placement of the subcutaneous needle electrodes before, during and after plasma administration. Specifically, the leads were placed in the standard Einthoven configuration for mice, ensuring that each mouse underwent the same procedure for accurate results. The lead configuration remained unchanged throughout the plasma infusion challenge. Here is a general description of the lead placement for mouse ECG:

– Lead I: This lead measures the potential difference between the right and left forelimbs (or arms). Electrodes are placed on both the right (positive) and left forelimbs (negative). This is shown in the first line of every ECG experiment.
– Lead II: This lead measures the potential difference between the right forelimb (negative) and the left hindlimb (positive). Electrodes are placed on the right forelimb and the left hindlimb. This is shown in the third line of every ECG experiment.
– Lead III: This lead measures the potential difference between the left forelimb (negative) and the left hindlimb (positive). Electrodes are placed on the left forelimb and the left hindlimb. This is shown in the second line of every ECG experiment.
– Ground Electrode: A ground electrode is placed on a neutral area, right hindlimb, to stabilize the signal and reduce noise.

Needle electrodes were connected via flexible cables to an amplifier (Micromed, Mogliano Veneto, Italy), then the ECG signal was recorded using System-Plus software (Micromed, Mogliano Veneto, Italy) and sampled at 256 Hz (16 bits) with band-pass filters between 1 and 70 Hz. All animal experiments were performed without prior knowledge of the origin of the plasma samples to ensure the integrity of the results.

### Data Analysis

Data were processed using GraphPad Prism (GraphPad Software, Inc.). Results are expressed as mean ± standard deviation (SD) or mean ± standard error of the mean (SEM) as appropriate. Statistical significance was determined using Student’s t-test or one-way ANOVA, depending on the nature of the data. A p-value of <0.05 was considered statistically significant.

## Results

### Study population characteristics

This study included a cohort of twenty patients diagnosed with metastatic breast cancer, each exhibiting diverse clinical profiles. The participants varied in age, cancer staging, and treatment regimens, which included a range of chemotherapeutic agents, targeted therapies, and supportive care, as detailed in Table 1. These patients were carefully selected based on specific inclusion criteria: diagnosis of early invasive or advanced stage breast cancer, prior treatment history, absence of chronic diseases. They also had normal ECG without BrS indicators^25^, no *SCN5A* variants, and no family history of BrS nor SCD. Additionally, six healthy individuals, all negative for BrS as confirmed by the ajmaline provocation test^25^, were included as control subjects for the immunoprecipitation and flow cytometry analyses.

**Table 1.** Clinical Characteristics of MBC patients.

| #ID | AGE RANGE | IMMUNEPHENOTYPE | CURRENT THERAPY | NaV 1.5 Ab |
| --- | --- | --- | --- | --- |
| 1 | 61-65 | BREAST CANCER | TD-M1+ CHT | + |
| 2 | 46-50 | BREAST CANCER | CHT | + |
| 3 | 41-45 | BREAST CANCER | TD-M1+ CHT | + |
| 4 | 71-75 | SKIN+ BREAST CANCER | MONOCLONAL ANTIBODY + CHT | + |
| 5 | 66-70 | BREAST CANCER | QUADRANTECTOMY | + |
| 6 | 56-60 | BREAST CANCER+COLON | 5-FLUOROURACIL + FOLINIC ACID + VITAMIN B12 | + |
| 7 | 51-55 | BREAST CANCER | CHT + MONOCLONAL ANTIBODY | + |
| 8 | 36-40 | BREAST CANCER | CHT | + |
| 9 | 61-65 | BREAST CANCER | TD-M1 | + |
| 10 | 61-65 | BREAST CANCER | MONOCLONAL ANTIBODY + CHT | + |
| 11 | 26-30 | BREAST CANCER | CHT | + |
| 12 | 86-90 | BREAST CANCER | MONOCLONAL ANTIBODY +CHT | + |
| 13 | 41-45 | BREAST CANCER | MONOCLONAL ANTIBODY A CHT | + |
| 14 | 51-55 | BREAST CANCER | CHT | + |
| 15 | 46-50 | BREAST CANCER | CYCLIN INHIBITORS + HT | + |
| 16 | 51-55 | BREAST CANCER | CHT | * |
| 17 | 41-45 | BREAST CANCER | CYCLIN INHIBITORS + HT | * |
| 18 | 56-60 | BREAST CANCER | CHT | * |
| 19 | 46-50 | BREAST CANCER | CHT | * |
| 20 | 41-45 | BREAST CANCER | CHT | - |

\* Unclear detection of NaV1.5 autoantibodies; signals were also present in untransfected HEK293A
cells, suggesting that these autoantibodies might target a protein with a similar molecular weight to
NaV1.5. CHT: Chemotherapy; TD-M1: Trastuzumab emtansine; HT: Hormonal Therapy.

### Detection of Anti-NaV1.5 Autoantibodies in metastatic breast cancer patients

HEK293A cells were engineered to express the adult NaV1.5 channel protein, confirmed by Western blot analysis with a commercial anti-NaV1.5 antibody, undetectable in mock-transfected cells (Figure 1A-B)^23^. Subsequently, the same membranes were incubated with plasma from MBC patients. Binding of autoantibodies to NaV1.5 was detected using a secondary anti-human IgG antibody (Figure 1A-B). Fifteen plasma samples from MBC patients showed a clear and detectable signal with the anti-human IgG, overlapping with the band previously stained with a commercial anti-NaV1.5 antibody (Figure 1B and Suppl. Fig. 1). One plasma sample did not show any detectable NaV1.5 autoantibodies. In four samples, a partially positive signal was also observed in control HEK cells, which do not express NaV1.5, suggesting that the autoantibodies may be binding also to other proteins with a similar molecular weight (Suppl. Fig. 9).

**Figure 1:**
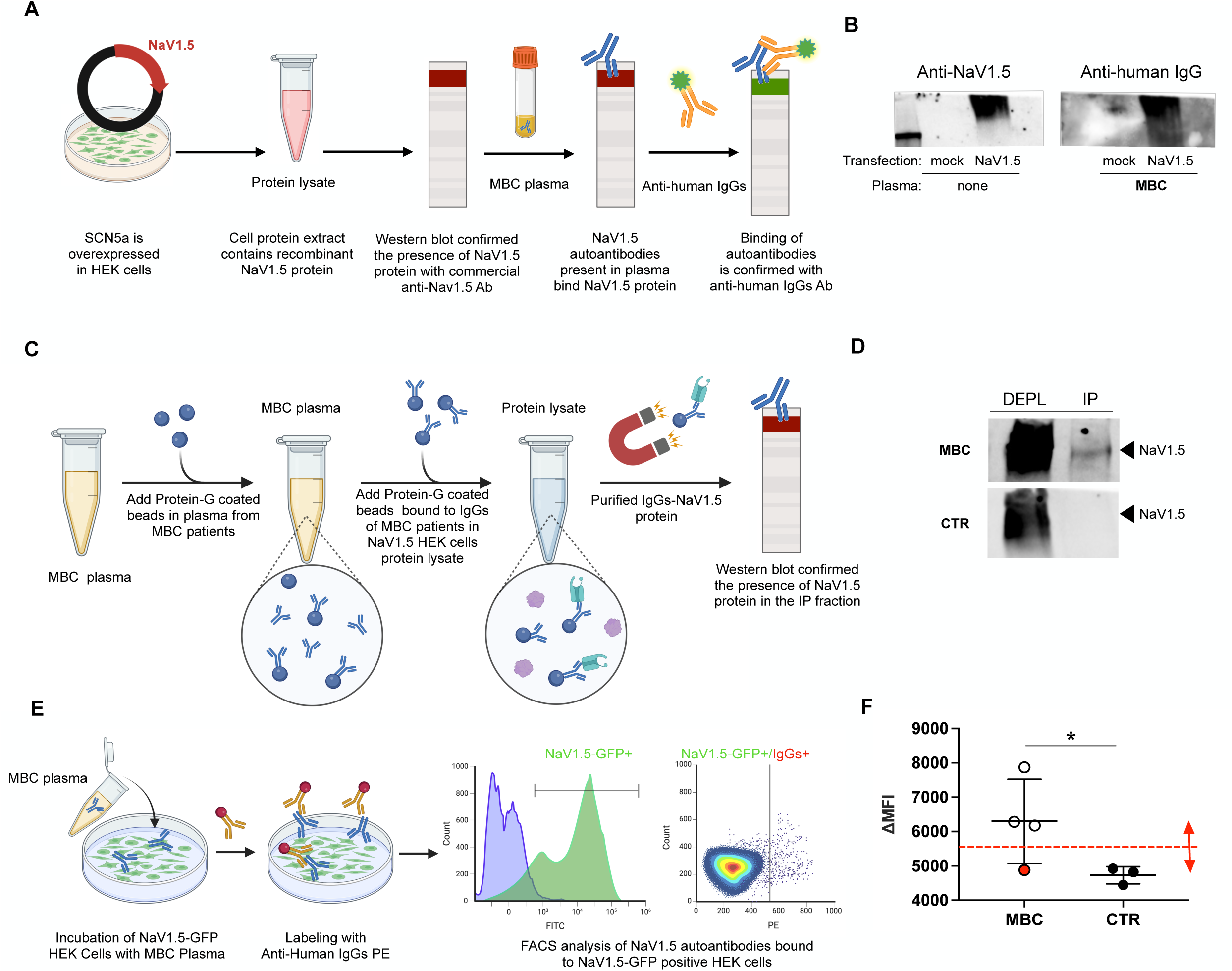
Detection of NaV1.5-Targeting Autoantibodies in Metastatic Breast Cancer (MBC) Plasma. (**A**) Schematic representation of the workflow for detecting anti-NaV1.5 autoantibodies in MBC plasma. HEK293A cells overexpressing NaV1.5 were lysed, and the proteins were resolved by SDS-PAGE. The membrane was first probed with a commercial anti-NaV1.5 antibody, followed by incubation with MBC plasma. Binding of plasma autoantibodies to NaV1.5 was detected using a secondary anti-human IgG antibody; (**B**) Representative Western blots. Left panel: The anti-NaV1.5 antibody confirms NaV1.5 protein expression in NaV1.5-transfected HEK293A cell lysates, but not in mock-transfected cells. Incubation with MBC plasma shows binding of autoantibodies to NaV1.5, detected by the anti-human IgG antibody; (**C**) Schematic of the immunoprecipitation workflow to detect anti-NaV1.5 autoantibodies in MBC plasma. Protein-G coated beads were used to pull down IgGs from MBC plasma, which were then incubated with NaV1.5-expressing HEK cell lysates to capture NaV1.5-targeting autoantibodies; (**D**) Western blot after immunoprecipitation (IP) shows NaV1.5 presence in immunoprecipitated samples, indicated by anti-NaV1.5 antibody staining. The analysis reveals a specific affinity of IgGs from MBC plasma for NaV1.5, in contrast to control plasma (CTR); (**E**) Schematic workflow for detecting NaV1.5-targeting autoantibodies in MBC plasma using flow cytometry. HEK cells expressing NaV1.5-GFP were incubated with MBC plasma, followed by staining with PE-conjugated anti-human IgG. Flow cytometry results show NaV1.5-GFP+ cells (left) and NaV1.5-GFP+/IgGs+ cells (right), indicating the presence of NaV1.5-specific autoantibodies in the plasma; (**F**) Quantification of flow cytometry data shows a significant increase in mean fluorescence intensity (ΔMFI) in MBC samples positive for NaV1.5 autoantibodies by Western blot, compared to controls (CTR) and MBC plasma without NaV IgG (red dot). The dotted red line suggests a tentative threshold for distinguishing positive samples from controls, though it requires further validation. This indicates higher levels of NaV1.5-targeting autoantibodies in the plasma of these MBC patients.

An immunoprecipitation experiment was then performed to confirm the specific binding of MBC IgGs to the NaV1.5 protein (Figure 1C). The results showed a positive NaV1.5 signal exclusively in the immunoprecipitated fraction when plasma from MBC patients was used. In contrast, no signal was detected when plasma from healthy controls was used (Figure 1D, Suppl. Fig. 2). The presence of anti-NaV1.5 IgGs was further assessed by flow cytometric analysis (Figure 1E-F, Suppl. Fig. 3). HEK293A cells expressing the NaV1.5-GFP channel were incubated with plasma from MBC patients and then stained with anti-human IgG. The results showed a significantly higher proportion of IgG-positive cells in the NaV1.5-overexpressing HEK293A cells incubated with MBC plasma compared to controls. Notably, the plasma sample from the MBC patient who tested negative for anti-NaV1.5 autoantibodies by Western blot showed results similar to the controls in the FACS analysis (Figure 1F, red dot).

### Effects of plasma of metastatic breast cancer patients on sodium currents in vitro

Prior to assessing the impact of MBC plasma on sodium currents, first was verified its non-toxic nature. MTT assays demonstrated that cell viability was preserved up to a 10% plasma concentration, although this concentration was sufficient to decrease the viability of the breast cell line (Suppl.Fig.4). Then, the effect of plasma from five MBC patients on I_Na_ was evaluated by incubating hiPS-CMs or transiently transfected HEK293A cells with 5% plasma with or without IgGs for 1 hour at 37°C and 5% CO_2_. Whole MBC plasma significantly reduced the current density in both cell models. Specifically, in hiPS-CMs, the TTX-sensitive peak current density measured at −20 mV was decreased by approximately 25% compared to untreated cells. In HEK293A cells, the peak current density of adult and neonatal NaV1.5 at −20 mV was reduced by approximately 34% and 45%, respectively. This inhibitory effect was completely abolished by the removal of IgG, as no significant changes in current density amplitude were observed when hiPS-CMs or HEK293A cells were incubated with IgG-depleted plasma from the same five MBC patients (Figure 2, Table 2). Interestingly, plasma from MBC patients had no effect on the voltage dependence of channel activation in hiPS-CMs. However, the activation curves of adult and neonatal NaV 1.5 expressed in HEK293A cells were significantly right shifted by approximately 3.2 mV and 4.9 mV, respectively. Removal of IgG from plasma abolished this effect only for the adult NaV1.5 isoform (Figure 2, Table 2). No effect of plasma on the steady-state inactivation curve was observed (data not shown). MBC plasma significantly reduced the TTX-sensitive sodium current in hiPS-CMs, with the effect being approximately 11.1% greater for the neonatal NaV1.5 isoform than for the adult isoform transiently expressed in HEK293A cells.

**Figure 2:**
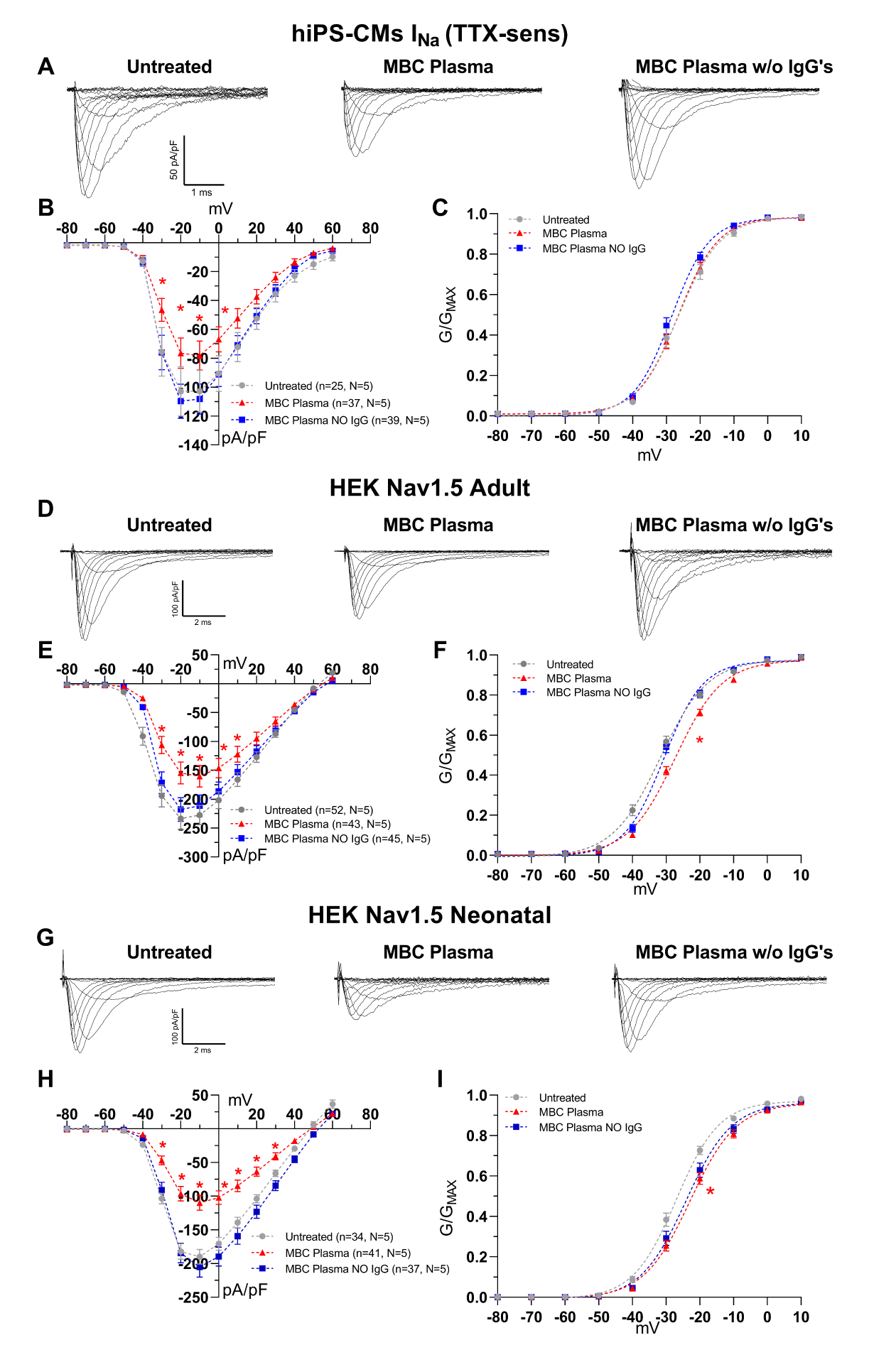
The effect of plasma from MBC patients on I_Na_. Gray circles, red triangles and blue squares represent untreated cells and cells incubated with plasma from MBC patients or the same plasma depleted of IgG, respectively. (**A**) Sample traces of TTX-sensitive I_Na_ recorded in untreated hiPS-CMs (left) or incubated with complete MBC plasma (center) or with the same plasma depleted of IgG (right). (**B**, **C**) I-V relationship and voltage dependence of channel activation in hiPS-CMs. (**D**) Sample traces of I_Na_ recorded in HEK293A transiently overexpressing the adult NaV1.5 isoform, untreated (left) or incubated with complete MBC plasma (center) or with the same plasma from which the IgGs were depleted (right). (**E**, **F**) I-V relationship of adult I_Na_ and voltage dependence of activation in HEK293A cells. (**G**) Sample traces of I_Na_ recorded in HEK293A transiently overexpressing the neonatal NaV1.5 isoform, untreated (left) or incubated with complete MBC plasma (center) or with the same plasma but without IgGs (right). (**H**, **I**) I-V relationship of neonatal I_Na_ and voltage dependence of neonatal NaV1.5 activation in untreated HEK293A cells. (n=number of cells, N=number of plasmas, *p<0.05 1 or 2-way ANOVA with Dunnet’s post-hoc Vs untreated and MBC plasma without IgGs).

**Table 2.** Effect of MBC plasma with or without IgG’s on sodium current in hiPS-CMs and on adult and neonatal Nav1.5 current in HEK cells.

| Current (cell line) | Parameters | Untreated | MBC Plasma | MBC Plasma w/o IgG's |
| --- | --- | --- | --- | --- |
| INa TTX-sensitive (hiPS-CMs) | Peak current density at -20 mV (pA/pF) | -102.9±17.1 | -76.9±10.5* | -109.8±11.8 |
|  | Activation V1/2 (mV) | -25.9±1.1 | -26.1±0.9 | -27.5±0.9 |
|  | Activation Slope (mV) | 5.1±0.3 | 5.1±0.2 | 4.6±0.2 |
|  | n (N) | 25 (5) | 37 (5) | 39 (5) |
| INa Adult (HEK293A) | Peak current density at -20 mV (pA/pF) | -233.6±18.2 | -154.6±19.0* | -217.8±20.5 |
|  | Activation V1/2 (mV) | -29.6±0.9 | -26.4±0.5* | -29.9±0.7 |
|  | Activation Slope (mV) | 5.1±0.3 | 6.9±0.2 | 5.4±0.2 |
|  | n (N) | 52 (5) | 43 (5) | 45 (5) |
| INa Neonatal (HEK293A) | Peak current density at -20 mV (pA/pF) | -182.2±11.5 | -100.2±11.1* | -184.5±14.6 |
|  | Activation V1/2 (mV) | -26.4±0.8 | -21.5±0.9* | -24.0±0.8 |
|  | Activation Slope (mV) | 6.1±0.2 | 6.6±0.3 | 6.0±0.2 |
|  | n (N) | 34 (5) | 41 (5) | 37 (5) |

n=number of cells, N=number of MBC plasmas, \*p<0.05 1 or 2-Way ANOVA with Dunnet's Post-hoc Vs untreated and MBC Plasma w/o IgG's

### Cardiac effects of metastatic breast cancer patient plasma injection in wild-type mice

Continuous ECG monitoring of mice expressing the wild-type isoform of aNaV1.5 channels that were intravenously administered plasma from three breast cancer patients who tested positive and one who tested negative for the presence of NaV1.5 autoantibodies, under general anesthesia showed that the MBC plasma cross-react with cardiac tissue (Suppl. Fig.2 and 6), and developed arrhythmic manifestations culminating in fatal electromechanical dissociation and death of the mouse following plasma exposure. The ECG recordings showed progressive AV conduction delay and significant ST elevation until the electromechanical dissociation and asystole leading to the death of the mouse. (Fig. 3 and Suppl. Fig. 6-9).

**Figure 3:**
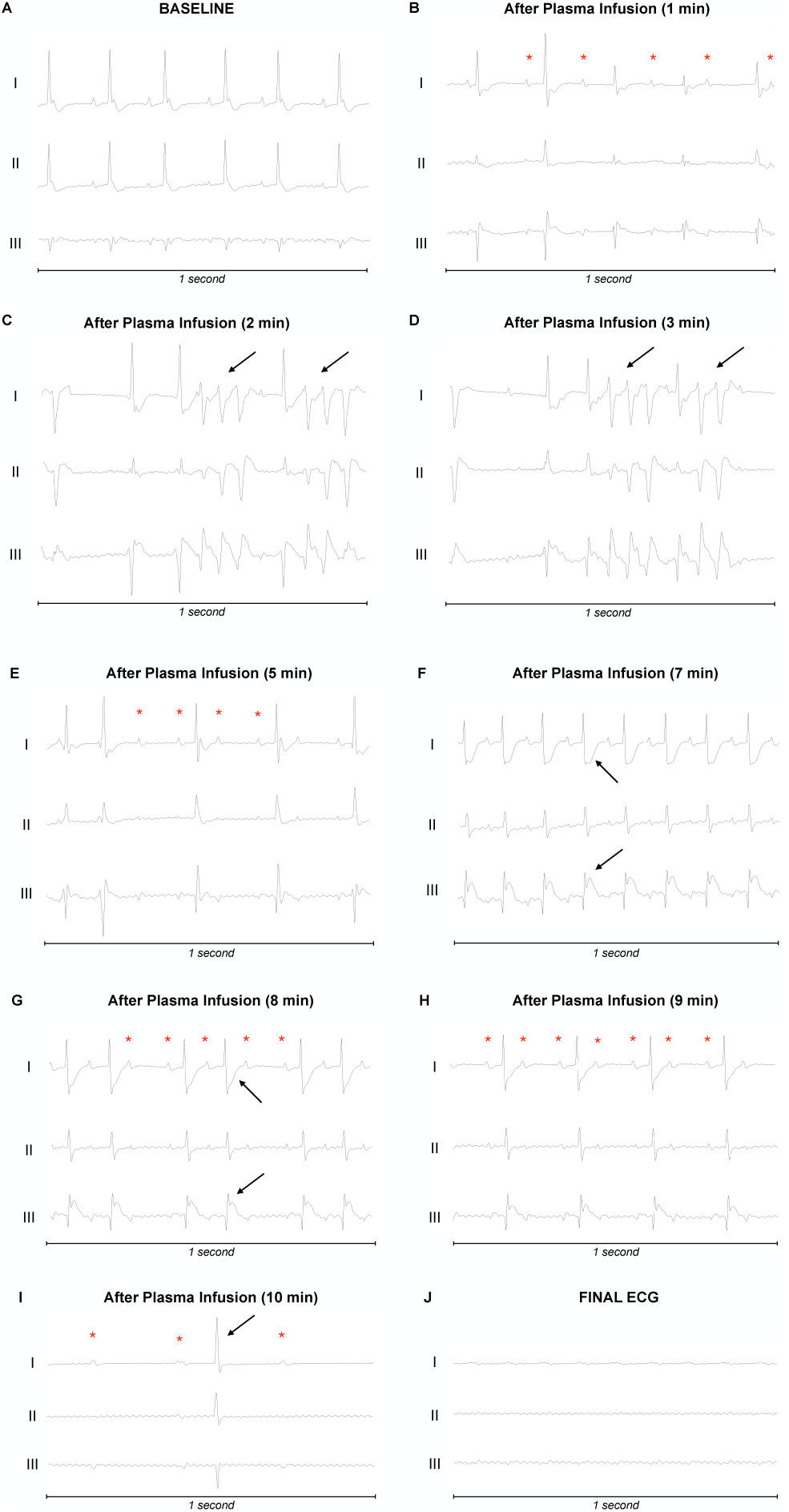
Electrocardiographic Response to MBC Plasma in a Mouse Model. Sequential ECG changes are shown in a mouse model following intravenous infusion of plasma derived from a breast cancer (MBC) patient. The panels illustrate a progression of arrhythmic events leading to fatal electromechanical dissociation and death. Each ECG recording spans 1 second. (**A**) Baseline ECG with normal sinus rhythm; (**B**) Progressive AV conduction delay (red asterisks indicate p-waves); (**C**-**D**) Non-sustained polymorphic ventricular tachycardia (black arrows), indicating a proarrhythmic effect; (**E**) Five minutes post-infusion, AV dissociation (red asterisk) and mild repolarization abnormalities appear, with significant ST elevation in lead III and reciprocal ST depression in lead I; (**F**) Seven minutes post-infusion, continued AV dissociation (red asterisks) and ST abnormalities (black arrow); (**G**-**H**) Persistent AV dissociation eight minutes post-infusion (red asterisks) with ongoing ST changes (black arrow); (**I**) Progressive atrioventricular block (red asterisk) and junctional escape rhythm (black arrow); (**J**) Electromechanical dissociation and asystole, culminating in the death of the mouse.

## Discussion

In this study, we show that autoantibodies, known to target the NaV1.5 channel overexpressed by breast cancer cells^22^, can cross-react with the cardiac isoform and potentially induce severe arrhythmias when injected into wild-type mice. These findings follow the recent discovery of pathogenic autoantibodies against the cardiac NaV1.5 channel in the plasma of patients with BrS^23^. Given the high homology among the isoforms of the NaV channel, we hypothesized that autoantibodies against any of the isoforms could cross-react with the cardiac isoform and thus possibly trigger arrhythmias. In this pilot study, we demonstrated that autoantibodies against NaV1.5 from metastatic breast cancer patients^22^, irrespective of patient age, cancer stage, or treatment regimen, significantly reduced inward sodium currents in HEK293A cells expressing the nNaV1.5 isoform (−45%). Importantly, these autoantibodies also cross-reacted with cells expressing the cardiac aNaV1.5 isoform (−34%), which differs from the neonatal isoform by only seven amino acids. Remarkably, injection of patient plasma into wild-type mice caused severe cardiac arrhythmias and ECG changes that were comparable to those observed after injection of BrS plasma under the same conditions^23^. This cross-reactivity can possibly trigger the fatal arrhythmias that are frequently observed in these cancer patients^2,5^. This is consistent with our previous findings that autoantibodies against cardiac NaV1.5 in BrS patients also inhibit other NaV isoforms, such as NaV1.4 in skeletal muscle^23^. The resemblance between the arrhythmic patterns observed in our study and those seen in BrS patients suggests that BrS might be a specific cardiac manifestation of a broader spectrum of immune-mediated channelopathies, which we propose to term “*lethal autoimmune channelopathies (LACs)*”. Although this perspective challenges conventional views and undoubtedly requires further investigation, it is crucial to note that these autoantibodies can induce fatal arrhythmias and a BrS-like ECG pattern when injected in healthy mice. This underscores their potential to trigger severe arrhythmias independent of pre-existing cardiac disease or structural abnormalities, pointing to the importance of considering autoimmune mechanisms in the management of arrhythmic disorders. This raises the possibility that pre-existing cardiac conditions or treatments, such as chemotherapy, could interact with these autoantibodies and further increase the risk of sudden cardiac death (SCD). These results also suggest that caution should be warranted when using sodium channel blockers in cancer patients. This is consistent with the results of clinical trials that have shown that cancer patients treated with common, non-specific sodium channel blockers have a higher mortality rate^28,29^. In addition, plasma from different BrS patients, each containing anti-NaV1.5 antibodies, was shown to produce varying effects in both in vitro and in vivo experiments, suggesting that the impact of these antibodies is dose-dependent^23^. This variation implies the existence of a threshold level of antibodies that determines the potential risk of arrhythmias. Importantly, this threshold may vary depending on pre-existing conditions affecting the heart, which could influence the severity of the arrhythmic effects. The development of a quantitative method, such as an ELISA immunoblot, to measure the presence and concentration of these autoantibodies could be particularly important in cancer patients to monitor their increased risk of SCD, especially in the initial phase of treatment and in the event of malignancy recurrence. Although we are still in the early stages of understanding the true prevalence of these *’Lethal Autoimmune Channelopathies*’, our findings suggest that we are only beginning to uncover the full extent of this condition, which appears to be driven by a common underlying mechanism across various diseases. Nonetheless, our findings support that regardless of the type of breast cancer, stage of disease, or treatment regimen, the presence of anti-NaV1.5 autoantibodies is consistent and exhibits significant cross-reactivity with cardiac NaV1.5 channels. While larger studies will be required, this initial observation seems to suggests that the potential for these autoantibodies to induce cardiac arrhythmias may be independent of specific clinical variables within this population. The focus on cross-reactivity rather than the type or stage of cancer highlights the urgency of considering these autoimmune interactions in the management of cancer patients, particularly when administering therapies that could exacerbate their arrhythmic potential. The detection of these autoantibodies in other epithelial cancers^30^, gliomas^31^, and lymphomas^32^, characterized by NaV1.5 protein expression is highly plausible and warrants further investigation^33^. In addition, these autoantibodies should also be searched in other diseases such as neurological disorders (e.g. epilepsy)^34^ and gastrointestinal diseases (e.g. inflammatory bowel disease and irritable bowel syndrome), which are known to be associated with alterations and *de novo* expression of sodium channels that could trigger an autoimmune response^35–37^.

### Study Limitations

This pilot study involved a limited number of patients. Nonetheless, the consistent findings across all samples provide robust support for our hypothesis, demonstrating a clear association between the presence of anti-NaV1.5 autoantibodies and the induction of Brugada Syndrome-like phenotypes. While further studies with larger cohorts are necessary to validate these findings and assess their broader applicability, we believe it is imperative to report these preliminary results given their significant clinical implications, particularly the potential need to reconsider the use of sodium blockers in cancer patients. Furthermore, while our study effectively utilized immunoblotting and electrophysiological assays for the detection and characterization of autoantibodies, these methods lack the quantitative precision of enzyme-linked immunosorbent assays (ELISA), which could provide deeper insights into the correlation between antibody levels and the severity of clinical manifestations.

## Conclusions

This study shows that anti-NaV1.5 autoantibodies in metastatic breast cancer patients can cross-react with the cardiac isoform, potentially leading to lethal arrhythmias. These findings suggest a possible mechanism behind the high rate of sudden cardiac death in this population. Screening tests for these autoantibodies, as well as targeted therapies to neutralize their effects, could reduce this risk in vulnerable patients. In addition, our mouse experiments suggest that these autoantibodies and their associated risks could be transmitted through blood transfusions. Further research is essential to fully understand the impact of lethal autoimmune channelopathies on the heart and to develop effective therapeutic interventions.

## Data Availability

All data produced in the present study are available upon reasonable request to the authors

## Acknowledgements

The authors thank the patients and family members for their help and participation in the study.

---

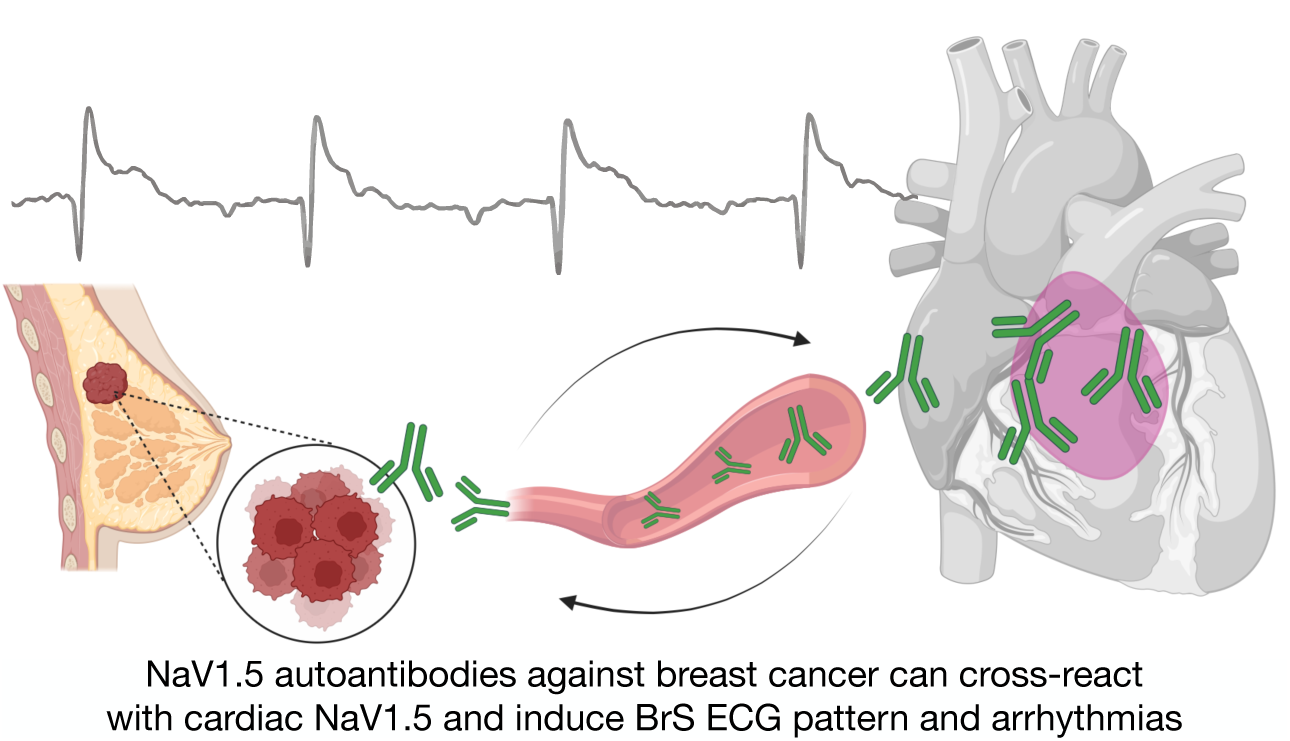

## Notes

### Competing Interest Statement

The authors have declared no competing interest.

### Funding Statement

This work received partial support from the Ricerca Corrente funding provided by the Italian Ministry of Health to IRCCS Policlinico San Donato and by IRCCS Policlinico San Donato own funds.

### Author Declarations

IRCCS Ospedale San Raffaele Ethics committee gave ethical approval for this work.

