## Supplementary Data for "Cardiac Cross-Reactivity of NaV Autoantibodies in Metastatic Breast Cancer: A Possible Trigger for Sudden Cardiac Death"

**Table of contents**


Supplementary Fig. 1. Pag 3

Supplementary Fig. 2. Pag 4

Supplementary Fig. 3. Pag 5

Supplementary Fig. 4. Pag 6

Supplementary Fig. 5. Pag 7

Supplementary Fig. 6. Pag 8

Supplementary Fig. 7. Pag 10

Supplementary Fig. 8. Pag 12

Supplementary Fig. 9. Pag 14

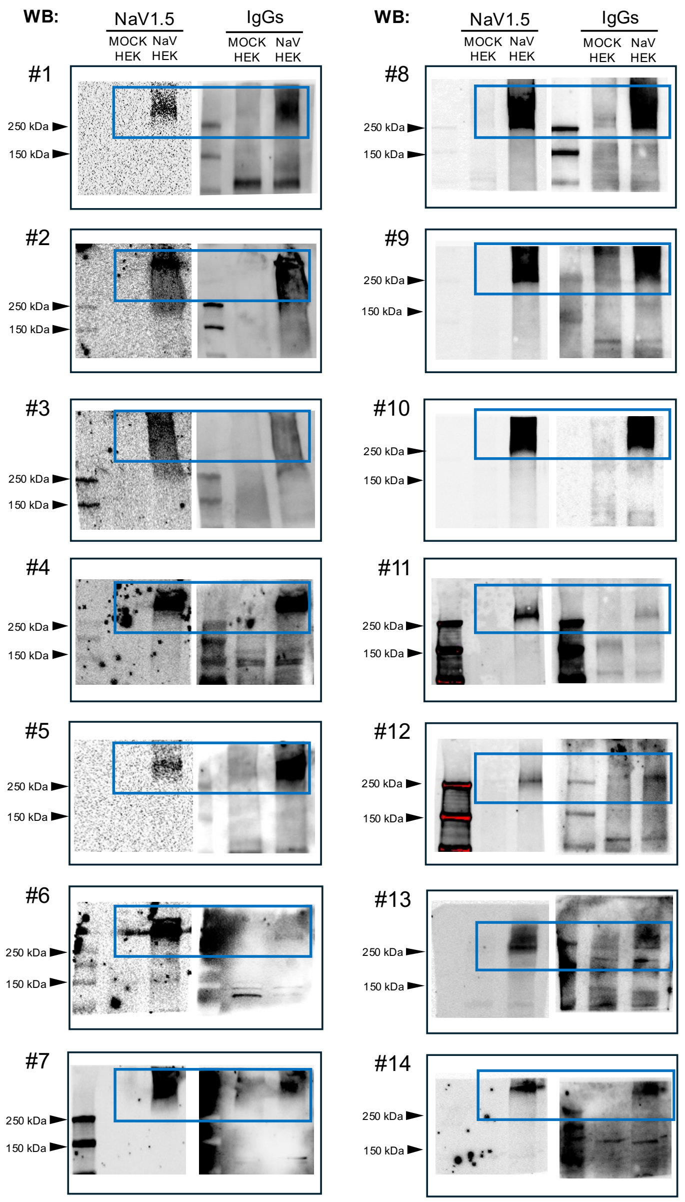


**Supplementary Fig. 1. Western Blot analysis to evaluate the presence of the NaV1.5 autoantibodies in the MBC study cohort.**

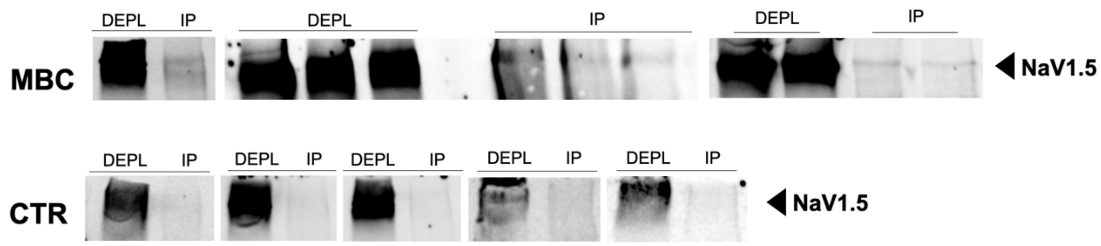


**Supplementary Fig. 2. Immunoprecipitation of NaV1.5 protein by NaV1.5-autoantibody.**

MBC (metastatic breast cancer patients' plasma) N=5; CTR (control plasma) N=6; IP (immunoprecipitated); DEPL (immunodepleted)

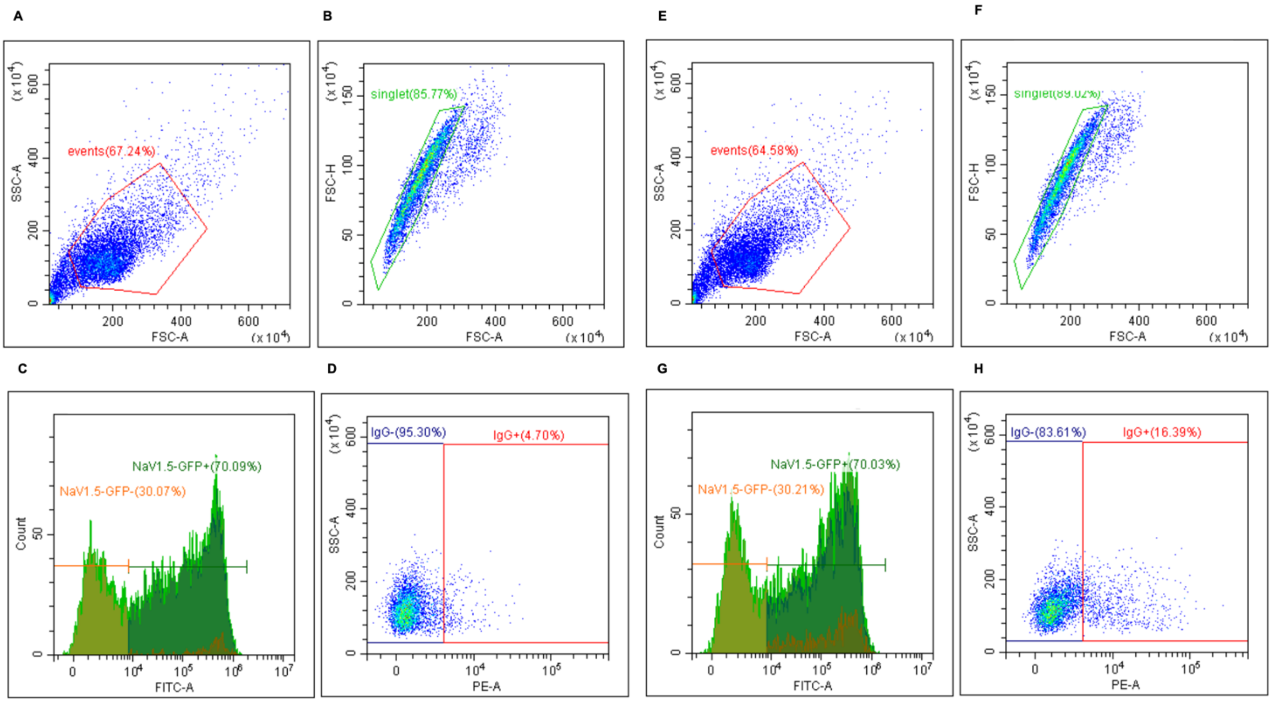


**Supplementary Fig. 3 Flow Cytometry gating strategy to detect Nav1.5+-IgG+**

Representative flow cytometry strategy gating for HEK293 cells. Flow cytometry data were gated based on forward scatter (FSC) versus side scatter (SSC) using a CytoflexS (**A, E**). Singlet cells were gated using FSC-A versus FSC-H (**B-F**). NaV1.5-GFP+ cells (**C, G**) were then selected to plot NaV1.5-GFP+/IgG+ populations (**D, H**).

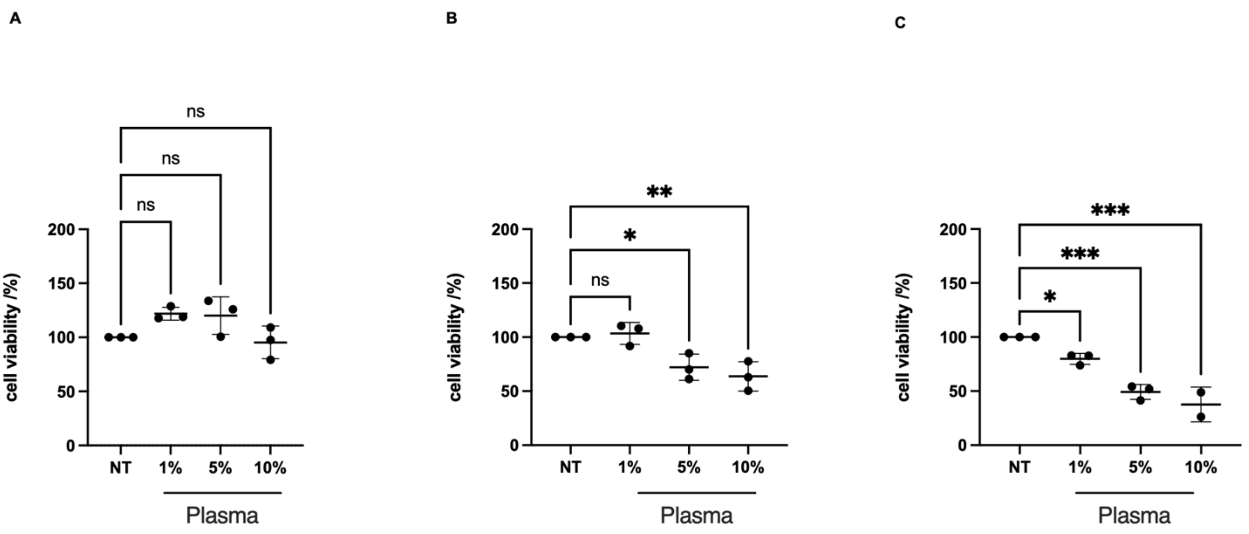


**Supplementary Fig. 4 Cytotoxic effect of MBC patient plasma *in-vitro*.** Cell viability post-treatment with different concentrations of MBC plasma was quantified using the MTT assay. (**A**) None of the plasma concentrations significantly impacted the viability of HEK293A cells; (**B**) except for the 1% concentration, all plasma concentrations significantly reduced cell viability in MDA-MB-231 cells; (**C**) all plasma concentrations significantly reduced cell viability in MCF-7 cells.

A B

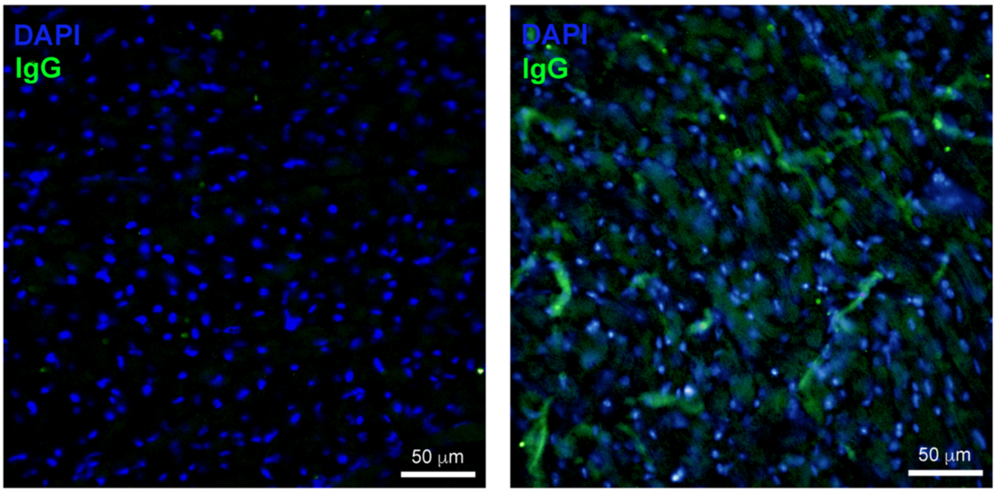


**Supplementary Figure 5. Presence of human IgGs in mice heart upon MBC plasma intravenous injection**

**(A)** Cardiac tissue from non-injected mice, (**B**) cardiac tissue from injected mice with MBC plasma.

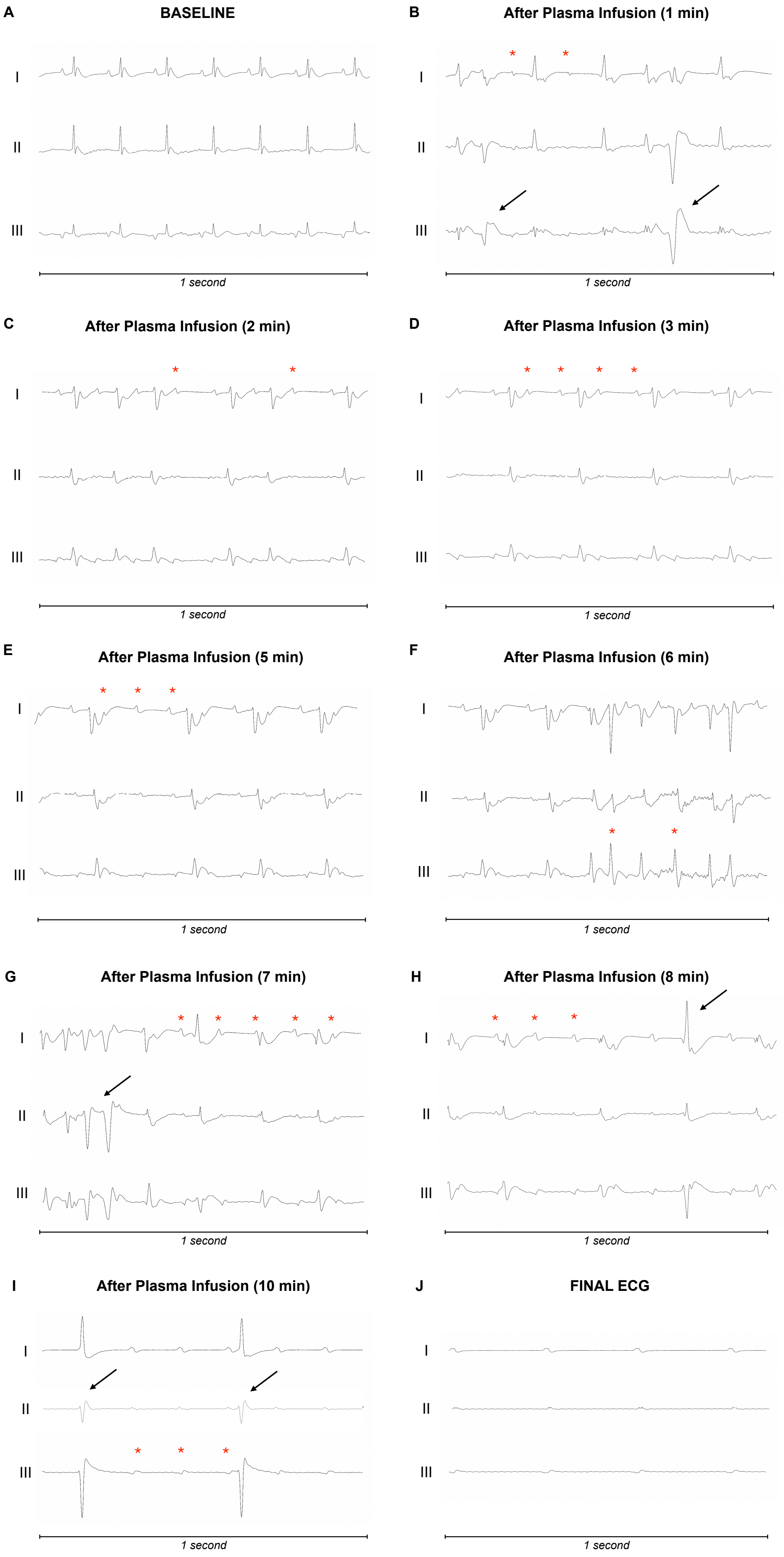
A
BASELINE
B
After Plasma Infusion (1 min)
C
After Plasma Infusion (2 min)
D
After Plasma Infusion (3 min)
E
After Plasma Infusion (5 min)
F
After Plasma Infusion (6 min)
G
After Plasma Infusion (7 min)
H
After Plasma Infusion (8 min)
I
After Plasma Infusion (10 min)
J
FINAL ECG
I
II
III
1 second

**Supplementary Figure 6. Experimental mouse #2 exposed to MBC patient plasma.** Sequential electrocardiographic changes in a mouse model following intravenous infusion of plasma from a breast cancer patient. Same ECG lead configuration as in Figure 3. (**A**) Baseline ECG with normal sinus rhythm; (**B**) Atrio-Ventricular (AV) conduction delay (red asterisk), and premature ventricular complexes occurrence (black arrows) after infusion; (**C**) QRS broadening and axis deviation, progressive atrio-ventricular (AV) conduction delay (red asterisk) and (**D**) 2:1 AV block, two and three minutes after infusion, respectively; (**E**) increasing degree of AV block (red asterisks indicate the P waves); (**F**) frequent premature complexes (red asterisks); (**G**) Seven minutes post-infusion, non-sustained polymorphic ventricular tachycardia (black arrow) with persistent AV dissociation (red asterisks); (**H**) Eight minutes after infusion, persistent complete AV dissociation (red asterisk) and junctional escape rhythm (black arrow) indicative of the severe sodium channel blockade; (**I**) junctional escape rhythm (black arrow), with AV dissociation; (**J**) Electromechanical dissociation and asystole leading to the death of the mouse.

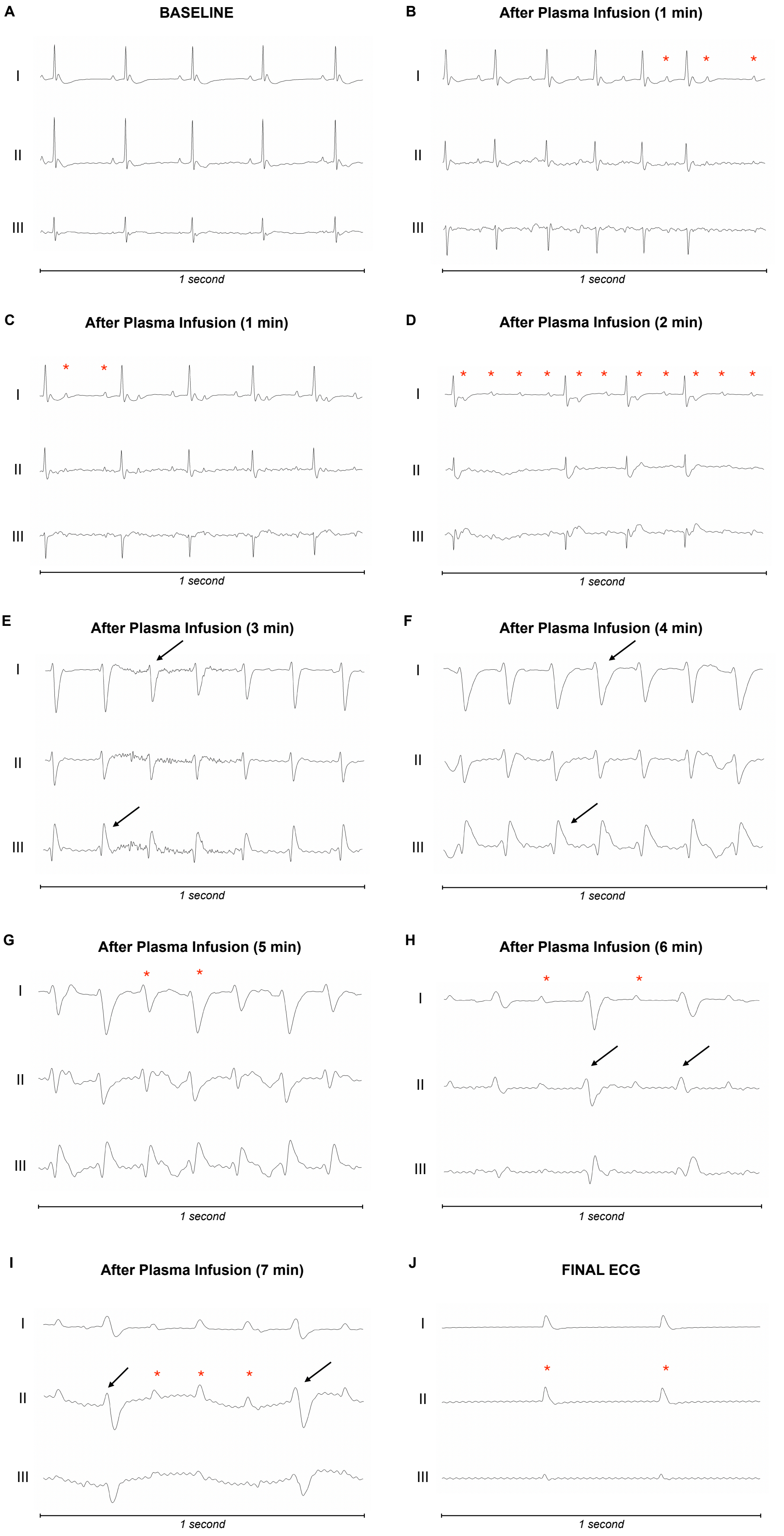
A
BASELINE
I
II
III
1 second
B
After Plasma Infusion (1 min)
I
II
III
1 second
C
After Plasma Infusion (1 min)
I
II
III
1 second
D
After Plasma Infusion (2 min)
I
II
III
1 second
E
After Plasma Infusion (3 min)
I
II
III
1 second
F
After Plasma Infusion (4 min)
I
II
III
1 second
G
After Plasma Infusion (5 min)
I
II
III
1 second
H
After Plasma Infusion (6 min)
I
II
III
1 second
I
After Plasma Infusion (7 min)
I
II
III
1 second
J
FINAL ECG
I
II
III
1 second

**Supplementary Figure 7. Experimental mouse #3 exposed to MBC patient plasma.** Sequential ECG changes in a mouse model following intravenous infusion of plasma from a breast cancer patient. Same ECG lead configuration as in Figure 3. (**A**) Baseline ECG with normal sinus rhythm; (**B**) atrio-ventricular conduction delay (red asterisk), after infusion; (**C**) one minute after infusion, 2:1 AV block; (**D**) progressive worsening of degree of AV block (red asterisk); (**E**) QRS broadening, axis deviation and repolarization abnormalities (black arrows); (**F**) severe QRS broadening and ST-segment elevation in lead III with reciprocal ST depression in lead I; (**G**) Five minutes post-infusion, broad QRS morphology with QRS amplitude alternans (red asterisks) indicating a severe sodium channel blockade; (**H**) Six minutes after infusion, persistent complete AV dissociation (red asterisk) and junctional escape rhythm (black arrow); (**I**) persistent AV block and junctional escape rhythm with wide QRS complexes (black arrow); (**J**) Electromechanical dissociation leading to the death of the mouse.

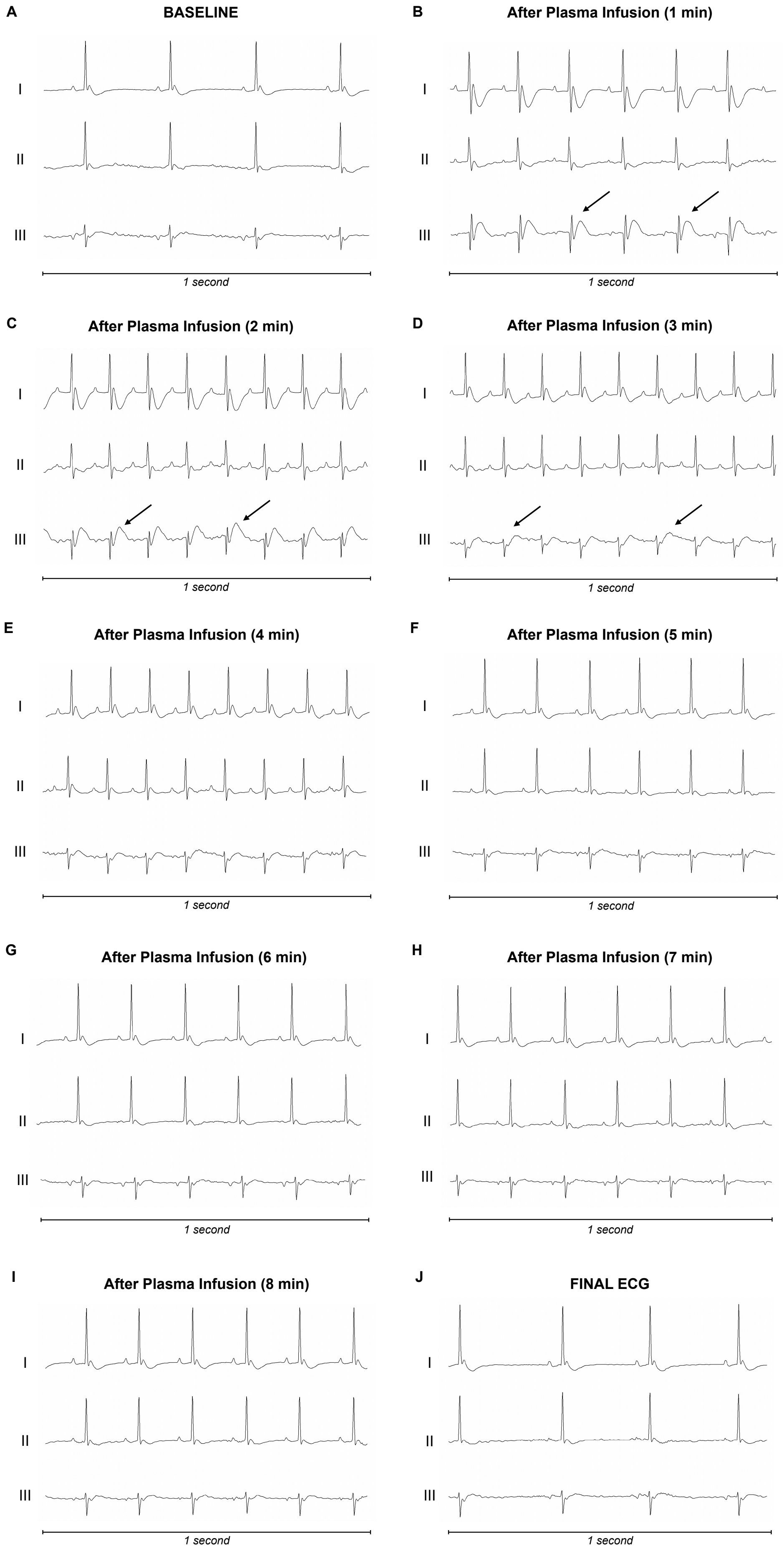
A
BASELINE
I
II
III
1 second
B
After Plasma Infusion (1 min)
I
II
III
1 second
C
After Plasma Infusion (2 min)
I
II
III
1 second
D
After Plasma Infusion (3 min)
I
II
III
1 second
E
After Plasma Infusion (4 min)
I
II
III
1 second
F
After Plasma Infusion (5 min)
I
II
III
1 second
G
After Plasma Infusion (6 min)
I
II
III
1 second
H
After Plasma Infusion (7 min)
I
II
III
1 second
I
After Plasma Infusion (8 min)
I
II
III
1 second
J
FINAL ECG
I
II
III
1 second

**Supplementary Figure 8. Experimental mouse #4 exposed to Antibodies-depleted MBC plasma**. This figure shows ECG recording in experimental mouse #4 exposed to the plasma from a MBC patient after antibody depletion. The plasma is from the same patient whose plasma, when inoculated in mouse #3, led to the ECG abnormalities, ST changes, AV block and death of the mouse. (**A**) Baseline ECG of this mouse shows normal sinus rhythm and electrical activity prior to plasma infusion; (**B**) one minute after infusion ST segment elevation in lead III and ST depression in lead I (black arrows); (**C-D**) progressive ST abnormalities attenuation at two and three minutes after plasma infusion (black arrows). The ECG consistently shows a normal sinus rhythm without any proarrhythmic effects and rapid ST-segment normalization throughout the observation time at four (**E**), five (**F**), six (**G**), seven (**H**) and eight (**I**) minutes after the infusion, contrasting with the progressive arrhythmogenic changes seen in mice subjected to the un-depleted MBC plasma. At the end of the experiment the mouse was still alive (**J**).

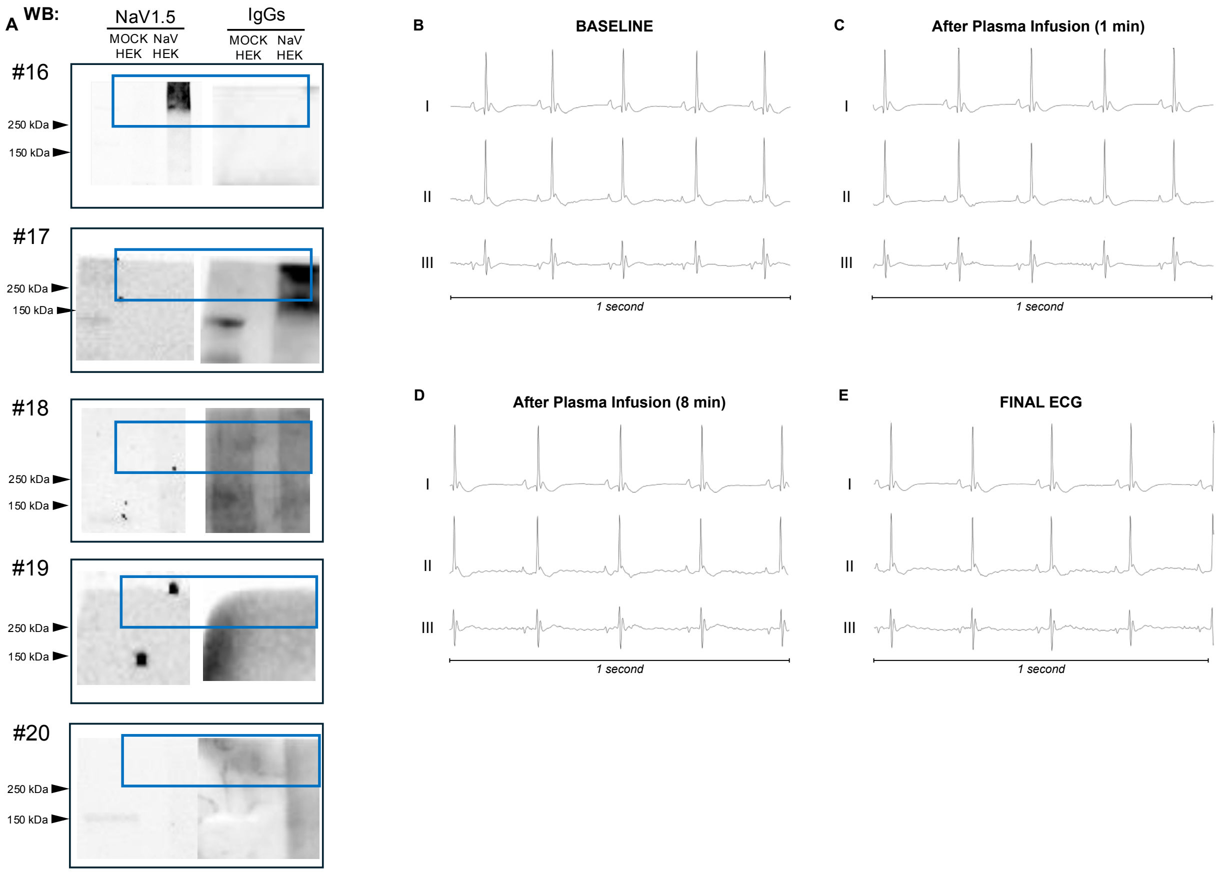


**Supplementary Figure 9. Analysis of NaV1.5 Autoantibody Presence and Cardiac Effects in MBC Patients with Unclear or Negative Staining**

(**A**) Western blot analysis evaluating the presence of NaV1.5 autoantibodies in plasma from five MBC patients with unclear or negative staining; (**B**) Baseline ECG of a mouse injected with plasma from an MBC patient who tested negative for NaV1.5 autoantibodies, showing normal sinus rhythm and electrical activity before plasma infusion; (**C-E**) ECG recordings at 1 minute, 8 minutes, and the final time point after plasma infusion, indicating no significant alterations throughout the experiment. The mouse remained alive at the end of the study.
